# Active vaccine safety surveillance via a scalable, integrated system in Australian pharmacies

**DOI:** 10.1101/2020.12.14.20248212

**Authors:** Sandra M Salter, Gurkamal Singh, Lisa Nissen, Kevin Trentino, Kevin Murray, Kenneth Lee, Benjamin Kop, Ian Peters, Alan Leeb

## Abstract

**Introduction:** Active vaccine safety surveillance will be critical to COVID-19 vaccine deployment. Pharmacists have been identified as potential immunisers in COVID-19 policies, yet there are no reported active surveillance systems operating in pharmacies. We integrated an established participant-centred active vaccine safety surveillance system with a cloud-based pharmacy immunisation-recording program. We measured adverse events following immunisation (AEFI) reported via the new surveillance system in pharmacies, and compared these to AEFI reported via an existing surveillance system in non-pharmacy sites (general practice and other clinics).

**Methods:** A prospective cohort study of individuals>10 years receiving influenza immunisations from 22 pharmacies and 90 non-pharmacy sites between March and October, 2020, in Australia. Active surveillance was conducted using SMS and smartphone technology, via an opt-out system. Multivariable logistic regression (including a subgroup analysis of participants over 65 years) was used to assess differences in proportions of AEFI between participants immunised in pharmacies compared to non-pharmacy sites, adjusting for confounders of age, sex, and influenza vaccine brand.

**Results:** Of 101,440 influenza immunisation participants (6,992 from pharmacies; 94,448 from non-pharmacy sites), 77,498 (76.4%) responded; 96.1% (n=74,448) within 24 hours. Overall, 4.8% (n=247) pharmacy participants reported an AEFI, compared with 6.0% (n=4,356) non-pharmacy participants (adjusted odds ratio: 0.87; 95% confidence interval: 0.76 to 0.99; p=0.039). Similar proportions of AEFIs were reported in pharmacy (5.8%; n=31) and non-pharmacy participants (6.0; n=1617) aged over 65 years (adjusted odds ratio: 0.94, 95% confidence interval: 0.65 to 1.35; p=0.725).

**Conclusion:** High and rapid response rates demonstrate good participant engagement with active surveillance in both pharmacy and non-pharmacy participants. Significantly fewer AEFIs reported after pharmacist immunisations compared to non-pharmacy immunisations, with no difference in older adults, suggests different cohorts attend pharmacy and non-pharmacy immunisers. The integrated pharmacy system is rapidly scalable across Australia with global potential.

**SUMMARY POINTS:** *What is already known?:*

- Many countries use post-licensure surveillance systems to monitor vaccine safety after a vaccine has been released onto the market.
- Passive surveillance systems operate in most countries but are limited by under- and/or biased reporting, and delayed detection of safety signals.
- Active surveillance systems have been reported for different sectors in Australia, the United States and Canada but there are no pharmacy-based active surveillance systems world-wide, and there is little evidence of rates of adverse events following immunisation (AEFI) in pharmacy.

*What are the new findings?:*

- We successfully linked two established platforms: an active vaccine safety surveillance system that integrates with national surveillance networks in Australia, with a cloud-based pharmacy immunisation-record system to develop an automated active vaccine safety surveillance system for pharmacies.
- Through active surveillance of 101,440 influenza immunisations between March and October, 2020 (6,992 from pharmacies, and 94,448 from non-pharmacy sites), fewer pharmacy participants reported any AEFI compared to non-pharmacy participants.

*What do the new findings imply?:*

- Pharmacists are safe immunisers who may capture patients not seen in general practice or other clinics.
- Our integrated pharmacy system is rapidly scalable, links with existing surveillance systems that integrate with the World Health Organization and has global potential.
- This study provides a proven infrastructure of crucial importance to maintain public safety, to promote confidence in vaccine safety, and to assist with COVID-19 vaccine uptake in a safe manner.

## INTRODUCTION

The novel SARS-CoV-2 virus and resulting COVID-19 pandemic has created health and economic crises of unprecedented proportions. Ultimately, only a safe and effective vaccine can mitigate the severe, disruptive consequences of COVID-19. At the time of writing, more than 200 vaccine candidates are under accelerated development – more than 50 of them under clinical evaluation (phase I, II or III human trials).[1]

Multiple vaccines are expected to be approved under individual government licensure rules and made available equitably through various initiatives including the COVAX Facility.[2] However, the unique and urgent need to deploy COVID-19 vaccines must be balanced against rapid vaccine development and limited safety data.[3-5]

Adverse events following immunisation (AEFI) can compromise public safety, undermine patient and immuniser confidence in any vaccine, and trigger markedly reduced rates of immunisation.[6] In context, vaccine safety surveillance extends beyond essential pharmacovigilance: it is critical to the deployment and continued uptake of a COVID-19 vaccine, as well as to ongoing success of existing immunisation programs.

Passive surveillance is the cornerstone of vaccine pharmacovigilance but relies on self-reporting of AEFIs by patients and healthcare professionals, and is limited by under- and/or biased reporting, and delayed detection of safety signals.[7,8] Active surveillance, by contrast, directly contacts patients after immunisation and can capture adverse event data from large numbers of vaccinees in near-real time.[9] Direct, participant-centred technological approaches, in particular those that are automated and integrated with larger networked systems,[8,10,11] offer broad scope and immediate promise for COVID-19 vaccine surveillance.

SmartVax is an established, participant-centred, automated active surveillance program that employs short message service (SMS) and smartphone technology,[8] and is a key element of Australia’s active vaccine safety monitoring system, AusVaxSafety.[12] This relationship facilitates sharing of Australia’s AEFI data with national agencies such as the National Centre for Immunisation Research and Surveillance (NCIRS) and the Therapeutic Goods Administration,[13] and with international agencies, notably the World Health Organization.[13]

Pharmacists have an established role in provision of influenza and other vaccines,[14,15] and in Australia, are also authorised to immunise patients in locations outside pharmacies (such as aged care facilities or schools).[16,17] Pharmacists have been identified as potential immunisers in COVID-19 vaccination policies,[18-20] yet, globally, there are no reported active surveillance systems operating in pharmacies. Given the important role pharmacists will play in COVID-19 immunisation, and their accessibility across cities, towns and remote regions, a broad-scale active surveillance system, with agility to adapt as new vaccines are approved, and capacity to operate across a range of geographical settings should be implemented. A system that is automated and linked directly to pharmacy software offers a simple, fail-safe option for surveillance, with little impact on pharmacist workload. MedAdvisor^®^ is a global, cloud-based, automated application for Pharmacy,[21] used widely in Australia. The MedAdvisor PlusOne^®^ platform is used by pharmacists to record immunisation encounters, and automatically reports immunisations to the Australian Immunisation Register (AIR).

In order to develop a participant-centred, automated active vaccine surveillance system for pharmacies, suitable for monitoring any vaccine delivered by pharmacists, we integrated SmartVax with MedAdvisor^®^. In this study we sought to test integration of the system through measurement of AEFI in pharmacies compared to existing non-pharmacy active surveillance sites, during the 2020 influenza immunisation season in Australia.

## METHODS

### Study design, participants and setting

We conducted a prospective cohort study of participants receiving immunisations from pharmacist and non-pharmacist providers in Western Australia between March and October 2020. Participants were consecutive individuals who self-selected to receive an influenza immunisation at any of 112 locations in Western Australia. This included 22 pharmacies recruited for the purposes of this study, and 90 non-pharmacy immunisation sites participating in ongoing active surveillance as part of usual practice.

The aforementioned 22 pharmacies were recruited for SmartVax integration in March 2020 via: direct email to all Western Australian (WA) community pharmacies using the MedAdvisor PlusOne^®^ platform to record immunisations; promotion through professional newsletters and Facebook pages; and via a media release by The University of Western Australia.[22] Site and pharmacist immuniser consent were obtained prior to enrolling each pharmacy into the study. All pharmacists were experienced in using the MedAdvisor PlusOne^®^ platform. Training on the SmartVax system and the study was provided to all pharmacists prior to data collection. All pharmacies received printed material to inform participants of the study consent process, with an explanatory poster for display in the immunisation consulting room.

Non-pharmacy sites (including general practice, university, community and local government clinics) participating in active surveillance through SmartVax have been previously described.[8,23]

SmartVax was used to actively monitor and record AEFIs between March and October 2020. Immunisations recorded in the MedAdvisor PlusOne^®^ system at each pharmacy were automatically batched and sent to SmartVax via a push application programming interface (API) each day. The API was used as a centralised mechanism to submit all records of vaccination encounters recorded across the MedAdvisor^®^ cloud application (for participating pharmacies) to the SmartVax server. Immunisation encounters recorded in the non-pharmacy sites were automatically extracted by SmartVax program at each site, each day.

SmartVax sent a series of automated SMS text messages to all participants three to five days after immunisation, to enquire if any AEFIs had been experienced. ‘Yes’ responders received a second SMS to enquire whether medical attendance had been sought, followed by a third SMS with a link to a short survey to identify the nature, duration and severity of all reactions (Figure 1). People who responded ‘No’ did not receive a second or third SMS.

**Figure 1.**
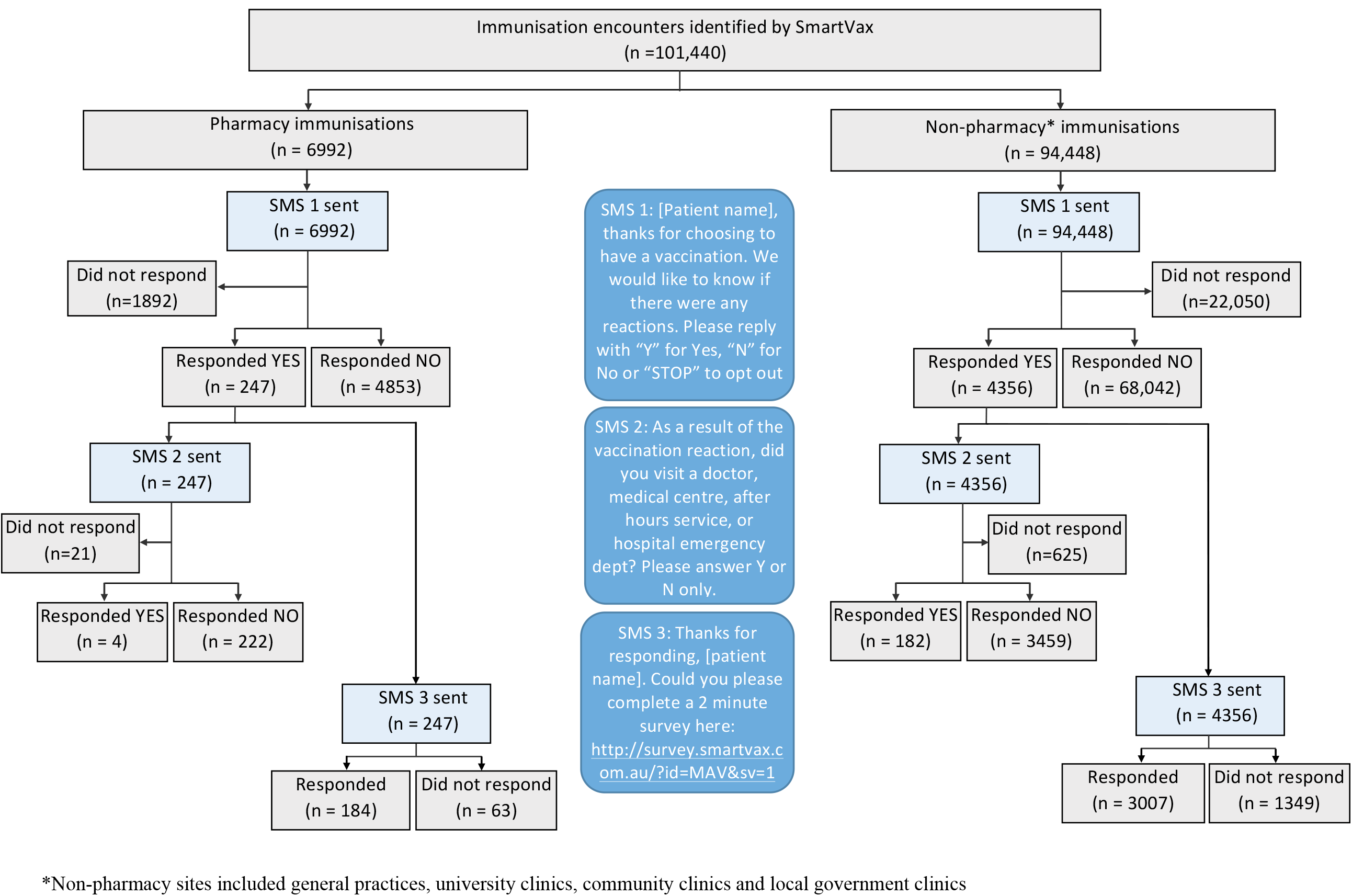
Flow diagram of patient identification for the study cohort. *Non-pharmacy sites included general practices, university clinics, community clinics and local government clinics

We also performed an audit of the integrated pharmacy system to determine the number of immunisation encounters recorded in the MedAdvisor PlusOne^®^ program that were 1) captured in the batch process, 2) made available for SmartVax surveillance, and 3) ultimately underwent surveillance.

Ethics approval for this study was obtained from The University of Western Australia Human Research Ethics Committee (RA/4/20/5907).

### Variables

The primary outcome of interest was any adverse event following immunisation (AEFI). Secondary outcomes included AEFIs resulting in medical attendance, and adverse event profile as reported in the survey (fever, pain, swelling, tired, irritable, sleep, rash, headache, vomiting, diarrhoea, and other). In addition to data collected on adverse events, SmartVax captured information on participant age, sex, and influenza vaccine brand. Pharmacists in WA are authorised to administer influenza vaccines to people 10 years and older, so we included age ≥ 10 years in our analyses. In order to compare adverse events reported by participants immunised in pharmacies to non-pharmacy sites, our exposure variable of interest was immunisation provider.

Data for the analysis were obtained from the SmartVax system, which includes participant demographic data, their responses to SMS messages, and where relevant, survey results.

### Statistical methods

Summary statistics for each immunisation location were provided and comparisons between pharmacy and non-pharmacy immunisations were made using the independent samples t-test (or Mann-Whitney test for non-normal data) for continuous variables, and chi-square tests for categorical variables. A comparison of participants reporting adverse events following influenza immunisation was made in unadjusted analysis using a chi-square test. Logistic regression was used to assess differences in proportions of adverse events between participants immunised in pharmacies and participants immunised at non-pharmacy sites, adjusting for confounders of age, sex, and influenza vaccine brand in a multivariable model. We conducted a sub-group analysis of participants 65 years and over. Only complete records (age, sex and vaccine brand) were included in the analyses. The significance level was set at 0.05.

All statistical analyses were conducted using R [version 3.5.3; The R Foundation for Statistical Computing]. Results are reported according to STROBE checklist for cohort studies.[24]

## RESULTS

We actively monitored 101,440 complete influenza immunisation encounters between 10 March 2020 and 17 October 2020 (6,992 from pharmacies, and 94,448 from non-pharmacy sites). A total 5,100 (72.9%) pharmacy participants and 72,398 (76.7%) non-pharmacy participants responded to SMS1, providing 77,498 immunisation encounters for analysis (Figure 1).

The mean (SD) age of participants was 51.5 (21.02) years, and 58··3% (n=45,186) were female. The most common influenza vaccines administered were FluQuadri 27.1% (n=27,491), Fluad Quad 26.6% (n=26,934), and Afluria Quad 10.3% (n=10,491). Overall, 96.1% (n=74,448) of responders replied within 24 hours of receiving SMS1, with 98.7% (n=76,485) responding within 72 hours. Of all responders, 5.9% (n=4,603) reported an adverse event following immunisation.

We present the characteristics of participants immunised in Table 1.

**Table 1.** Characteristics of influenza immunisation encounters by location.

|  | Location |  | p-value |
| --- | --- | --- | --- |
|  | Pharmacy<br>(n=5,100) | Non-pharmacy<br>(n=72,398) |  |
| Sex, female (%) | 3016 (59.1) | 42170 (58.2) | 0.218 |
| Age, years (mean (SD)) | 46.52 (17.02) | 51.90 (21.23) | <0.001 |
| Age, years (minimum, maximum) | 10,99 | 10,101 |  |
| Age group, years |  |  | <0.001 |
| <18 | 371 (7.3) | 7314 (10.1) |  |
| 18 - 64 | 4196 (82.3) | 38042 (52.5) |  |
| >= 65 | 533 (10.5) | 27042 (37.4) |  |
| Vaccine brand (%) |  |  | <0.001 |
| Afluria Quad | 1974 (38.7) | 8517 (11.8) |  |
| Fluad Quad | 514 (10.1) | 26420 (36.5) |  |
| Fluarix Tetra | 1119 (21.9) | 6862 (9.5) |  |
| FluQuadri | 914 (17.9) | 26578 (36.7) |  |
| Fluzone High-Dose | 0 (0.0) | 2 (0.0) |  |
| Influvac Tetra | 567 (11.1) | 1416 (2.0) |  |
| Vaxigrip Tetra | 12 (0.2) | 2600 (3.6) |  |
| SMS response time, minutes (median [IQR])* | 13 [3,72] | 14 [4,70] | 0.006 |
| Responses received to SMS1 within 24 hours (%) | 4939 (96.8) | 69549 (96.1) | 0.006 |

Values are count (proportion), mean (SD) or median (IQR) as indicated. P-values are
calculated from the chi-square test for categorical variables and the independent samples t-test for continuous variables. \* The p-value for SMS response time was calculated using the Mann-Whitney Wilcoxon Test.

Participants receiving influenza vaccinations at pharmacies had a lower mean age (46.5 vs. 51.9 years; p<0.001), with 10.5% (n=533) aged 65 years and over compared with 37.4% (n=27,042) at non-pharmacy sites. Pharmacists were more likely to administer Afluria Quad^®^ (Seqirus) and Fluarix Tetra^®^ (GlaxoSmithKline), while non-pharmacist immunisers were more likely to administer Fluad Quad^®^ (Seqirus) and FluQuadri^®^ (Sanofi-Aventis) (p<0.001). Response times to the first SMS were similar between groups, with a median (IQR) of 13 minutes (3 to 72 minutes) for pharmacy, and 14 minutes (4 to 70 minutes) for non-pharmacy vaccinees.

### Rate of any adverse event

The unadjusted proportion of any adverse event differed significantly between participants immunised at pharmacies and those immunised elsewhere. Of those immunised at pharmacies, 4.8% (n=247) reported an adverse event, compared with 6.0% (n=4,356) for those immunised at non-pharmacy sites (unadjusted odds ratio = 0.80, 95% confidence interval 0.70 to 0.91; p=0.001). After adjusting for age, sex, and vaccine brand, participants immunised at pharmacies reported fewer adverse events, compared to participants immunised in non-pharmacy sites (odds ratio (OR)=0.87, 95% confidence interval (95% CI): 0.76 to 0.99, p=0.039).

### Adverse events resulting in medical attendance

Participants reporting any adverse event were sent a second SMS asking if they visited a doctor, medical centre, after-hours service, or hospital emergency department as a result of their vaccination reaction. We excluded 646 (0.8%) encounters from this analysis, as they did not respond to the second SMS.

Of participants immunised at pharmacies, 0.1% (n=4) reported seeking medical care following an adverse event compared with 0.3% (n=182) of participants immunised at non-pharmacy sites (p=0.021). After adjusting for age, sex, and vaccine brand, participants immunised at pharmacies reported less adverse events for which medical attendance was sought, compared to participants immunised at non-pharmacy sites (OR=0.35, 95% CI: 0.13 to 0.97, p=0.042).

### Adverse event profile

The most common adverse events reported after immunisation in pharmacies were pain (2.0%; n=104), tiredness (1.9%; n=95), and headache (1.7%; n=88). The most common adverse events reported after immunisation in non-pharmacy sites were pain (2.3%; n=1660), tiredness (1.9%; n=1362), and swelling (1.5%; n=1121), Figure 2. A total of 1.4% (n=68) pharmacy and 1.2% (n=778) non-pharmacy participants selected ‘other’ AEFI, and included a text description of the reaction. The majority of these reactions were described as aches, dizziness, muscle and joint pain, cold symptoms, sore throat, and nausea; Supplementary Tables 1 and 2. Figure 3 presents odds ratios for adverse event profile after adjusting for age, sex, and vaccine brand, comparing participants immunised at pharmacies to those immunised at non-pharmacy sites.

### 65 years and over sub-analysis

Of participants aged 65 years and over, 5.8% (n=31) immunised at pharmacies reported any adverse event compared with 6.0% (n=1617) for those immunised at non-pharmacy sites (OR= 0.97, 95% CI: 0.67 to 1.40; p=0.875). After adjusting for age, sex and vaccine brand, participants immunised at pharmacies reported similar proportions of any adverse events when compared to participants immunised at non-pharmacy sites (OR=0.94, 95% CI: 0.65 to 1.35, p=0.725). No participants 65 years and over who were immunised at pharmacies reported seeking medical care following an adverse event, compared with 0.3% (n=72) of participants immunised at non-pharmacy sites.

### Audit of the integrated pharmacy system

Of 11,008 immunisations given in the pharmacies, 10.0% (n=1106) chose not to participate in surveillance, and 71.0% (n=7821) were captured in the API batch process. Of these, 95.6% (n=7475) were available for SmartVax surveillance, and 92.4% (n=7230) were sent SMS1. We identified in July 2020 that 20.1% (n=2081) records for participants without a recorded sex had not transferred via the API. This was rectified and subsequently 100% of records (including records without sex) were successfully transferred via the API (Figure 4).

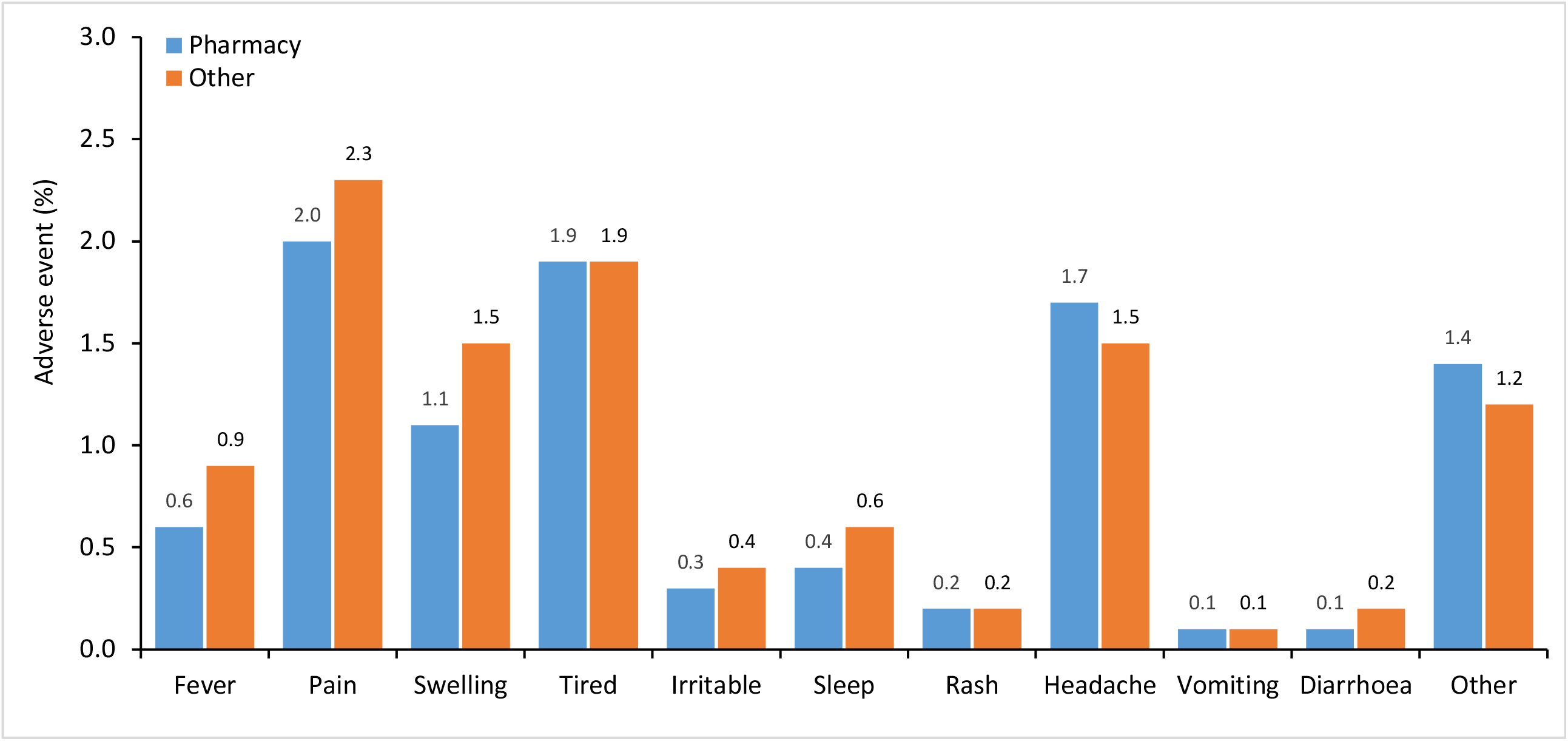

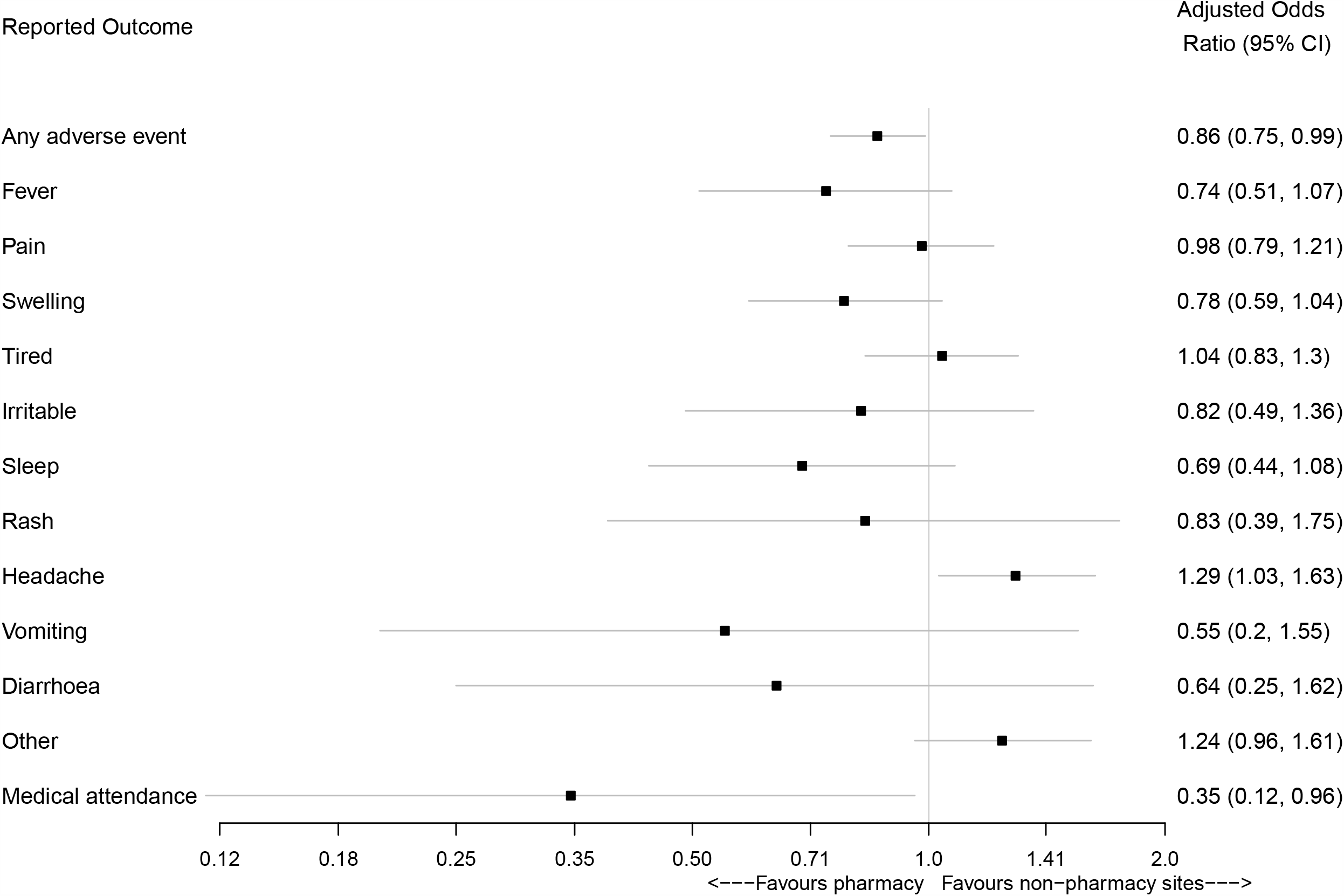

**Figure 4:**
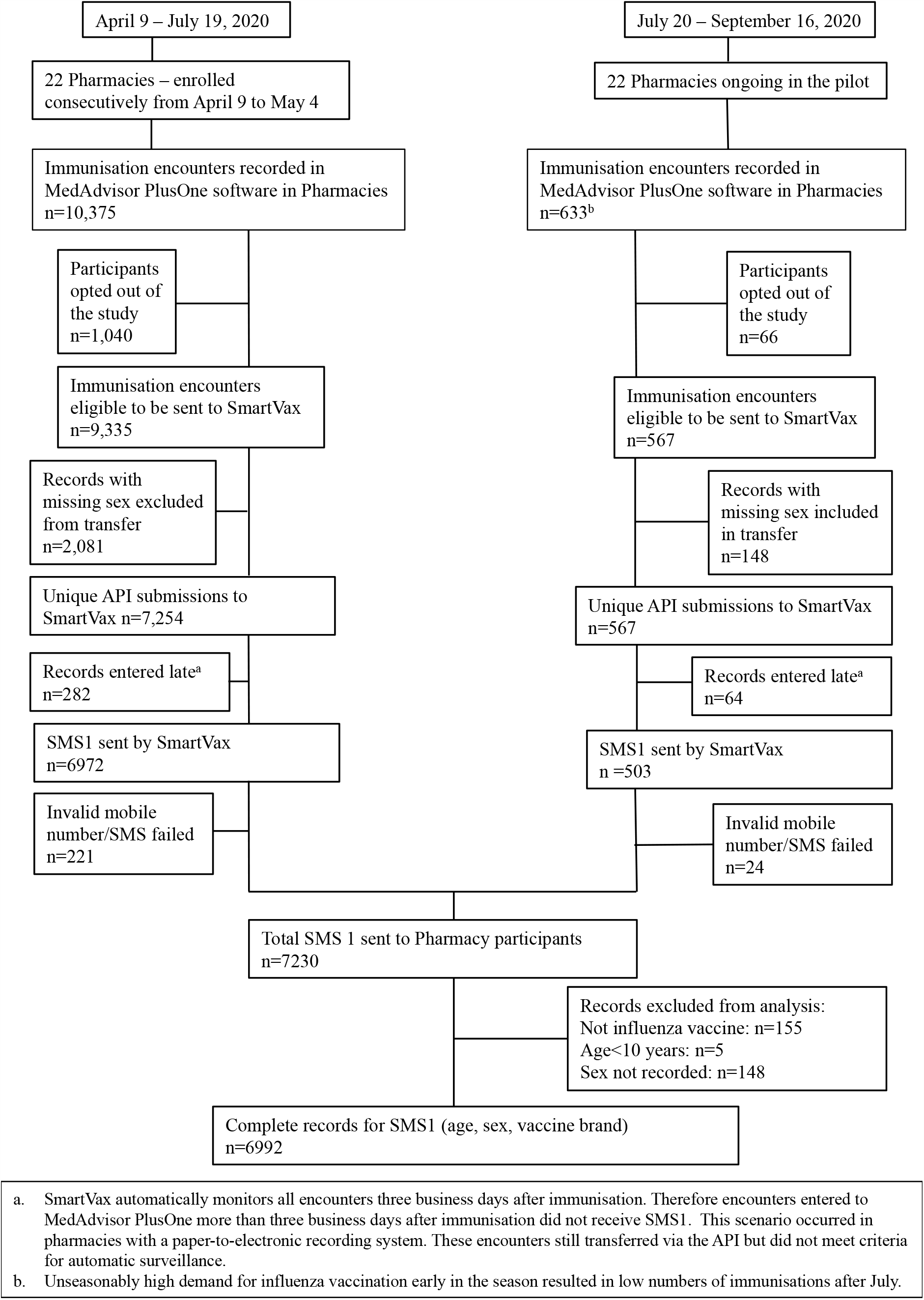
Audit of data flow in the integrated pharmacy active vaccine safety surveillance system. a. SmartVax automatically monitors all encounters three business days after immunisation. Therefore encounters entered to MedAdvisor PlusOne more than three business days after immunisation did not receive SMS1. This scenario occurred in pharmacies with a paper-to-electronic recording system. These encounters still transferred via the API but did not meet criteria for automatic surveillance. b. Unseasonably high demand for influenza vaccination early in the season resulted in low numbers of immunisations after July.

## DISCUSSION

Active surveillance systems, particularly those that integrate new technologies with current vaccine surveillance systems,[3,25] will be vital to maintain public health and confidence, in what may be the world’s biggest immunisation program in history: global immunisation against COVID-19. This study is the first in the world to implement and evaluate active surveillance of adverse events following immunisation (AEFI) in pharmacies. Through integration of established active vaccine surveillance and cloud-based pharmacy software systems, we developed a robust, rapidly scalable, automated, user-friendly, large scale AEFI monitoring system.

There was a high (73%) and rapid response to active surveillance following influenza immunisation in pharmacies (median 13 minutes; 96% within 24 hours); similar to responses observed in non-pharmacy sites, both in this study and previously.[8,23] While the profile of adverse events reported after pharmacy-based immunisations was similar to that reported after immunisation in non-pharmacy sites, there were significantly fewer adverse events overall reported from participants immunised in pharmacies compared to those immunised in non-pharmacy sites, including general practice and other clinics. After adjusting for age, gender, and vaccine brand this difference was still significant, albeit clinically small. As participants immunised in pharmacies were significantly younger than those in non-pharmacy sites, it is possible that more complex or sicker participants visited non-pharmacy immunisers. However, there was no difference in overall adverse events observed in our subanalysis of the over 65 years age group, whom we might expect to have more comorbidities or be sicker. Notably, a surge in demand for early influenza vaccination in 2020, means we did not capture all participants immunised in participating pharmacies, as our integrated system was activated a month after pharmacy vaccinations began. This could have included sicker patients of any age. The relationship with the immuniser, anticipation of AEFI, or sense of obligation to report even minor reactions may be different for people attending pharmacies and non-pharmacy sites, but we did not measure this. Furthermore, each site can and does immunise patients opportunistically. Such immunisation may, by default, affect the cohort attending and their perception of AEFIs: people opportunistically immunised in non-pharmacy sites (including general practice) may perceive themselves to be sicker and be more inclined to report AEFIs, although we did not measure the proportion of booked vs. opportunistic immunisations. Regardless, given response rates and time to respond were similar between groups, the reason for the observed difference and whether it is a true effect remains unclear. Even if the difference is true, it may simply represent different cohorts being immunised in different sites: this in itself is of benefit as it indicates pharmacies may capture a different set of people who may otherwise not seek immunisation. Importantly, engagement with the integrated pharmacy system was observed in 5100 participants (aged 10-99 years) across 22 pharmacies, illustrating that age was not a barrier to pharmacy vaccination, nor the technology used in the system.

This study has the potential to change practice immediately. There are two cloud-based platforms used in Australia to record vaccination encounters, each of which has the capacity to automatically upload the immunisation record to the Australian Immunisation Register (AIR),[26] yet until now there was no structured surveillance system in pharmacy and no mechanism to identify reactions once patients left the pharmacy. Through this study, active vaccine safety surveillance is now instantly scalable to thousands of pharmacies in Australia and can monitor patients previously out of reach. Furthermore, there is potential for global scalability given the cloud-based pharmacy software integrated in this study, MedAdvisor, has an international presence. This may be particularly useful for pharmacies in countries where there does not appear to be published evidence of ongoing active surveillance systems, for example, the United Kingdom.[27]

Beyond this, proposed changes to legislation requiring all vaccination providers to report vaccines given to the AIR[28] means all pharmacy platforms will also have capacity to report to the SmartVax API, extending coverage of surveillance. Significant economies of scale are achieved through the pharmacy integration: technical support, program updates and modifications to surveillance parameters can be completed efficiently in either component of the integrated system and quickly activated at all participating sites. The system also has potential for growth: in research, to link our surveillance data to national or state based administrative datasets for long term surveillance and comparison of long term outcomes against early reported AEFI and SARS-CoV-2 infection; in surveillance, to automatically extract additional data such as medication lists, for incorporation in surveillance analyses.

Connection through the integration to AusVaxSafety, the principal Australian AEFI active surveillance system, means pharmacy AEFI can be included in publicly reported surveillance data, ensuring open and transparent monitoring. In context of high social media-led disinformation campaigns about vaccine safety,[29] open reporting promotes public confidence in the vaccine and the surveillance system, and can mitigate anti-vaccination campaigns.[30] While provider recommendations drive immunisation,[6] public and provider belief that regulatory systems are robust will be critical to achieve high COVID-19 immunisation rates required for herd immunity.[31]

Rapid engagement of a trained immunisation workforce with access to active vaccine safety surveillance will be crucial for COVID-19 vaccine deployment. Pharmacists have been recognised as qualified immunisers in many countries for more than ten years.[15] Two systematic reviews have demonstrated pharmacists increase vaccination coverage when compared to traditional (non-pharmacist) providers alone.[32,33] Several factors specific to pharmacies including convenience, accessibility, extended opening hours, and the ability to obtain a vaccination without prior appointment have been shown to increase immunisation rates across the world.[14,15,32,33] It is likely pharmacies capture patients who would otherwise avoid immunisation,[14] but this is not driven by cost: non-pharmacist immunisers in Australia are partly funded by the Australian Government to provide immunisation services, whereas pharmacists receive no remuneration. Yet patients who could access partly or fully subsidised vaccines through a general practitioner choose to obtain, and pay for, their immunisations from pharmacies.[14,15,34]

While COVID-19 vaccines will be free in Australia,[18] several factors beyond cost and safety surveillance could influence uptake. During the first wave of COVID-19 infections when caseload was high, the Australian Government launched a National Health Plan, to protect both the public and the primary care workforce.[35] This resulted in a significant shift to telehealth consultations, with high uptake in general practice,[35] although pharmacies remained open as ‘essential services’ to the public to provide medicines and primary care.[36] The active COVID-19 caseload at the time of vaccine deployment may not be known,[18] nor the impact this might have on the available immunisation workforce. While pharmacists in Australia are not expected to deliver COVID-19 immunisations during initial deployment, they can still influence vaccine uptake through education and advocacy,[15,32] deliver a range of routine vaccinations and provide critical surge capacity[37] including for COVID-19 vaccines, and surveillance in the early event non-pharmacist immunisers are overloaded or under lockdown. Pharmacists can capture patients who are fearful of encountering acutely infectious individuals in clinic waiting rooms,[15] and should be considered, with active surveillance on board, as early potential vaccinators for Australia. Finally, suitably trained hospital-based pharmacists may also contribute via immunisation of health care workers and others in hospitals.

The integrated system is not without limitations. Our active surveillance integration requires Internet connectivity for the immunisation provider and mobile network connectivity for the consumer. This may pose an issue for the global population, where Internet access is estimated to be approximately 63%, with approximately 67% subscribed to mobile services,[38] but is less of an issue in developed countries with broad coverage.

Approximately 10% of participants in this study chose not to participate in active surveillance and did not receive SMS1. This may have been a feature of not wishing to participate in a research project, rather than wishing to avoid monitoring, and is expected to be different in the rollout of a COVID-19 vaccine. It is also important to note causality assessment as per the WHO criteria is an essential component of any vaccine safety surveillance program.[39] This is not achievable using our integrated system (only a temporal association is observed), nor is it intended: the purpose is to provide near-real time surveillance, with a known denominator, so that early vaccine safety signals can be identified. Causality assessments are within the scope of the wider network, including AusVaxSafety and the NCIRS.

A key strength of this study was our examination of data flow, to ensure all immunisation records transferred in the integrated pharmacy system. To the best of our knowledge this is the only audit of data flow in an active surveillance system. We identified three key issues. Firstly, the API did not identify records without sex recorded, and as a result, 21% of immunisations were not automatically monitored in the first half of this study. This issue was identified, the system was updated, and all records subsequently transferred. Future iterations of the system should enforce recording of all patient data during the consent process. Secondly, a small proportion of pharmacists used a paper-based system to record vaccination encounters, and subsequently transposed those records to the electronic system. This may have been a stop-gap measure during the unprecedented demand period of early COVID-19 activity, or it may represent usual practice. Records transferred more than three business days after immunisation were not automatically monitored, and the reason for the paper system was not elucidated. Thirdly, 3% of participants did not have a valid mobile phone number recorded and also were not monitored. Attention should be given to identify whether participants own a mobile phone, and if so, to accuracy in data entry.

## CONCLUSION

We are on the cusp of a new period in immunisation. The crucial next step is to leverage proven technology to enable broad scale vaccine deployment of any COVID-19 vaccines that are approved for use, with a trained workforce enabled with active surveillance systems. We have developed an integrated active vaccine safety surveillance system that is immediately scalable to thousands of pharmacies in Australia and potentially globally. Lower proportions of adverse events following immunisation in pharmacies compared to non-pharmacy sites demonstrates pharmacies are a safe destination for immunisation, and may capture people who would not otherwise obtain an immunisation. With an integrated system that facilitates both reporting of the immunisation to the Australian Immunisation Register, as well as links to national vaccine safety surveillance, pharmacists in Australia can contribute actively and safely to all immunisation programs.

## Data Availability

Individual participant data that underlie the results reported in this article, after de-identification (text, tables, figures, and supplementary material) are available upon reasonable request.
Proposals for data should be directed to Sandra Salter via ORCID: https://orcid.org/0000-0002-5840-6797.
To gain access to data, users will need to sign a data access agreement. Reuse is permitted under the following conditions: meta-analyses of adverse events following immunisation in pharmacies. 

## ACKNOWLEDGMENTS

The study was supported by a research grant provided by the J.M. O’Hara Research Fund of the Pharmaceutical Society of Western Australia.

We are extremely grateful for the tremendous in-kind support provided by SmartVax and MedAdvisor International and for their unending commitment and tireless efforts to conduct and complete the study in the year 2020 – a year of significant disruption on a global scale. We would also like to thank Ms Dani Li, Mr Craig Schnuriger, Mr Weyn Ong, Mr Daniel Smith, Ms Karin Orlemann, and Ms Danae Perry, for their continued efforts and contributions.

We are extremely grateful for the tremendous in-kind support provided by SmartVax and MedAdvisor International and for their unending commitment and tireless efforts to conduct and complete the study in the year 2020 – a year of significant disruption on a global scale. We would like to thank Ms Dani Li, Mr Craig Schnuriger, Mr Weyn Ong, Mr Daniel Smith, Ms Karin Orlemann, Ms Danae Perry, Ms Rachel Nesaraj and Mr Nicolas Bchara for their continued efforts and contributions.

